# Respiratory Viral Infection Patterns in Hospitalised Children before and after COVID-19 in Hong Kong

**DOI:** 10.1101/2024.06.07.24308528

**Authors:** Jason Chun Sang Pun, Kin Pong Tao, Stacy Lok Yam, Kam Lun Hon, Paul Kay Sheung Chan, Albert Martin Li, Renee Wan Yi Chan

**Affiliations:** Department of Paediatrics, Faculty of Medicine, The Chinese University of Hong Kong (CUHK); Hong Kong Hub of Paediatric Excellence, CUHK; Laboratory for Paediatric Respiratory Research, Li Ka Shing Institute of Health Sciences, CUHK; Department of Microbiology, Faculty of Medicine, CUHK; SH Ho Research Centre for Infectious Diseases Faculty of Medicine, CUHK

**Keywords:** Co-infection of respiratory viruses, paediatric respiratory virus epidemiology, age-specific responses, post-COVID-19

## Abstract

In the wake of Hong Kong’s zero-COVID policy, this study comprehensively analyses the epidemiological shift in respiratory viruses among hospitalized pediatric patients. The research leverages a unique natural experiment created by the policy’s stringent measures, which led to a significant reduction in virus circulation from 2020 to early 2023. The study highlights two distinct periods: pre-COVID-19 and post-mask mandate. We used pediatric hospitalization records from January 2015 to December 2019 and March 2023 to February 2024 to reveal a notable rebound in respiratory viruses. The age-stratified analysis indicated a shift in virus susceptibility. The odds ratio of having a co-infection was significantly increased in hospitalized children aged <1 to 12 years old during the post-COVID-19 mask mandate. Moreover, the adenovirus infection in younger children was more prominent, while RSV expanded its prevalence to older children aged>6 years old and raised health concerns. The study underscores the potential long-term impacts of interrupted virus exposure on children’s immune development and the need for vigilant monitoring of respiratory virus trends. It calls for further research to elucidate the causal relationships between SARS-CoV-2 exposure, subsequent respiratory virus susceptibility, and the implications for paediatric health in the post-pandemic era.

**Highlights:**

- Children (4.37 ± 0.05 years old) hospitalized after COVID-19 and tested for respiratory viruses were significantly older than those (3.49 ± 0.03 years old) before COVID-19.
- The odds ratio of having a co-infection was significantly increased in hospitalized children aged <1 to 12 years old during the post-COVID-19 era.
- In the post-COVID-19 era, the adenovirus infection in younger children was more prominent while RSV expanded its prevalence to older children aged>6 years old

## Introduction

Hong Kong, a city that stands out for its implementation of a zero-COVID policy (1), has provided a unique setting for our study. This policy, which included strict border control, mandatory quarantine, widespread testing, contact tracing, and universal masking, led to a distinct period in Hong Kong with minimal respiratory virus circulation, except for SARS-CoV-2. This unique environment served as a natural experiment to assess whether the disappearance of respiratory viruses would be associated with a change in respiratory virus epidemiology when the population resumed their normal routine after the removal of the mask mandate on March 1, 2023.

Our focus on children is crucial due to their ongoing lung and immunity development. Exposure to viruses at different age windows can influence the development of immune memory and, consequently, their responses to the same virus compared to those regularly exposed during childhood. The COVID-19 pandemic can potentially alter the intensity of respiratory virus exposure in certain age groups. Therefore, an age-stratified analysis is vital to determine if a specific virus is more closely linked to the vulnerability of a particular age group. This underscores the potential adverse effects on children’s health, the increased burden on the healthcare system, and the necessity for adjustments in hospitalization and treatment strategies.

## Methods

To investigate this, our study analyzed the prevalence patterns of respiratory viruses in hospitalized pediatric patients in Hong Kong, leveraging the well-defined periods of stringent ‘zero-COVID’ policies. Our study extracted the paediatric hospitalization records of those below 18 years old at the Prince of Wales Hospital from January 2015 to February 2024. We included subjects who had respiratory symptoms and gave specimens from their upper airways to test for various respiratory viruses during admission. The array of viruses includes adenovirus (AdV), influenza A virus A (IAV), influenza B virus B (IBV), parainfluenza viruses (PIV) 1, 2, 3, 4, respiratory syncytial virus (RSV), and coupled enterovirus and rhinovirus (EV/RV) were included in this study. We classified the era as ‘pre-COVID-19 era’ from January 2015 to December 2019, ‘COVID-19 lockdown’ from January 2020 to February 2023, and ‘post-COVID-19 mask mandate period’ from March 2023 to February 2024. The prevalence of individual viruses was defined by the number of positive detections among all the virology tests performed, with all samples retained as the denominator to address the fluctuations in sample size over the study period. Samples with uncertain results were excluded from the analysis. The paediatric subjects were stratified into five groups, including infants (<1y), toddlers (1y to <3y), preschool (3y to <6y), grade-schoolers (6y to <12y), and adolescents (12y to <18y).

## Results & Discussion

19,378 and 6,001 respiratory specimens with a complete panel of virus PCR tests in the pre-COVID-19 and post-COVID-19 mask mandate periods, respectively, were included in this study (**Table 1**). Before the pandemic, EV/RV, influenza, and RSV represented the major burdens amongst the paediatric admissions. During the COVID-19 lockdown, most respiratory virus infections were abolished while sporadic spikes of EV/RV and a minimal occurrence of AdV remained (**Figure 1**). A gradual rebound of viral infections was observed in the post-COVID-19 mask mandate. Moreover, AdV and EV/RV have shown a notably increased prevalence (**Table 1** Upper panel).

**Table 1:**
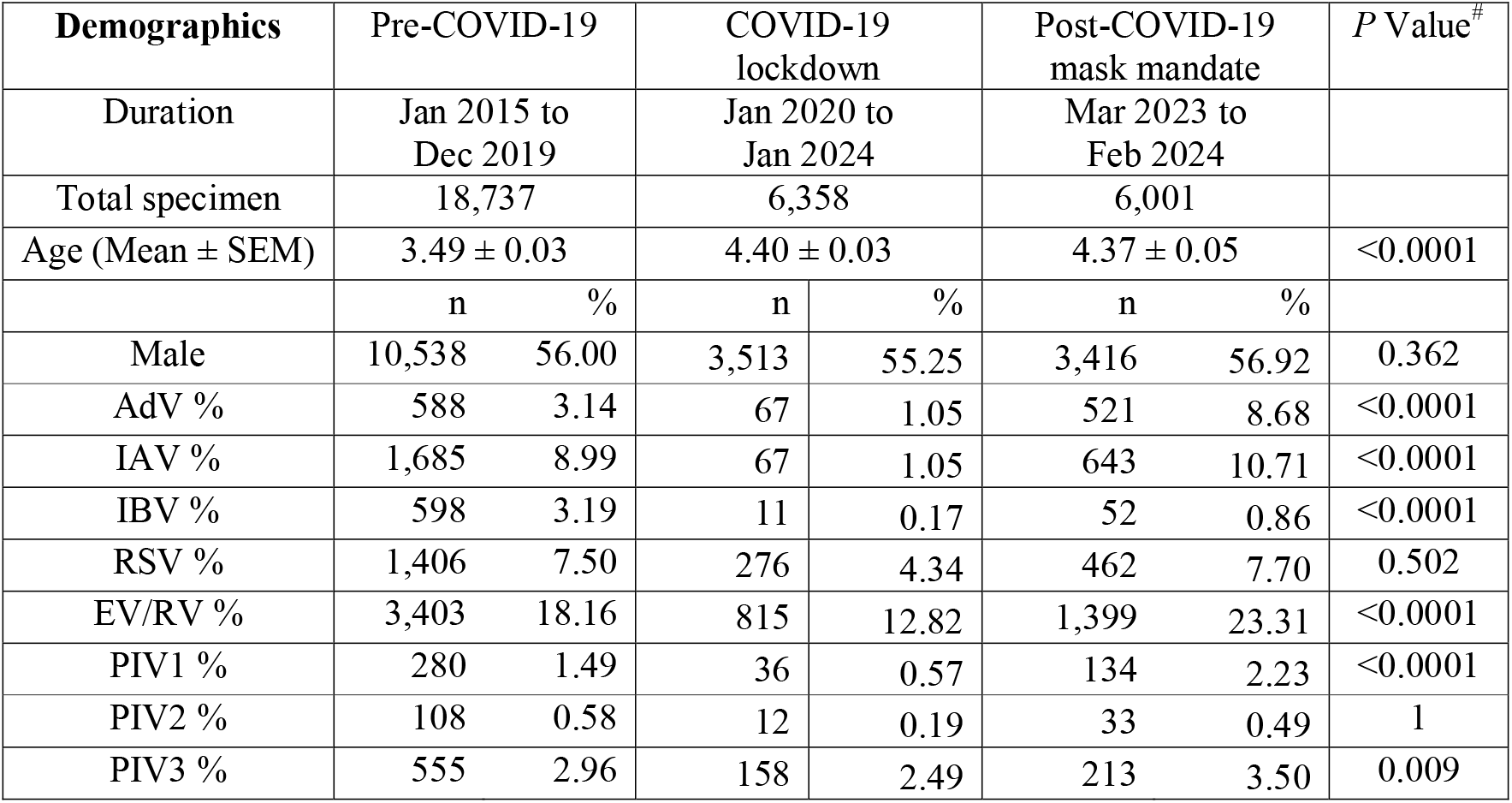
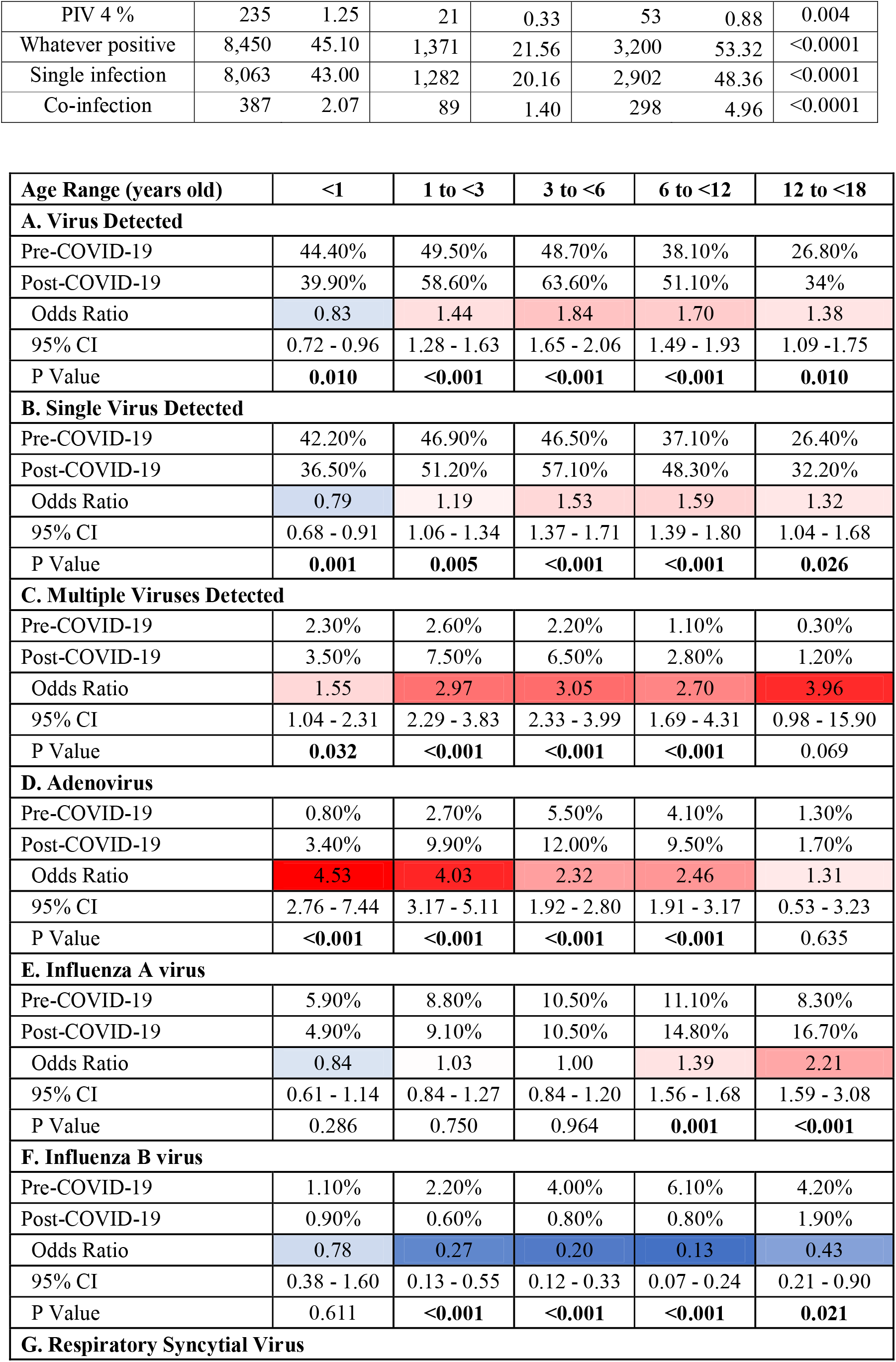
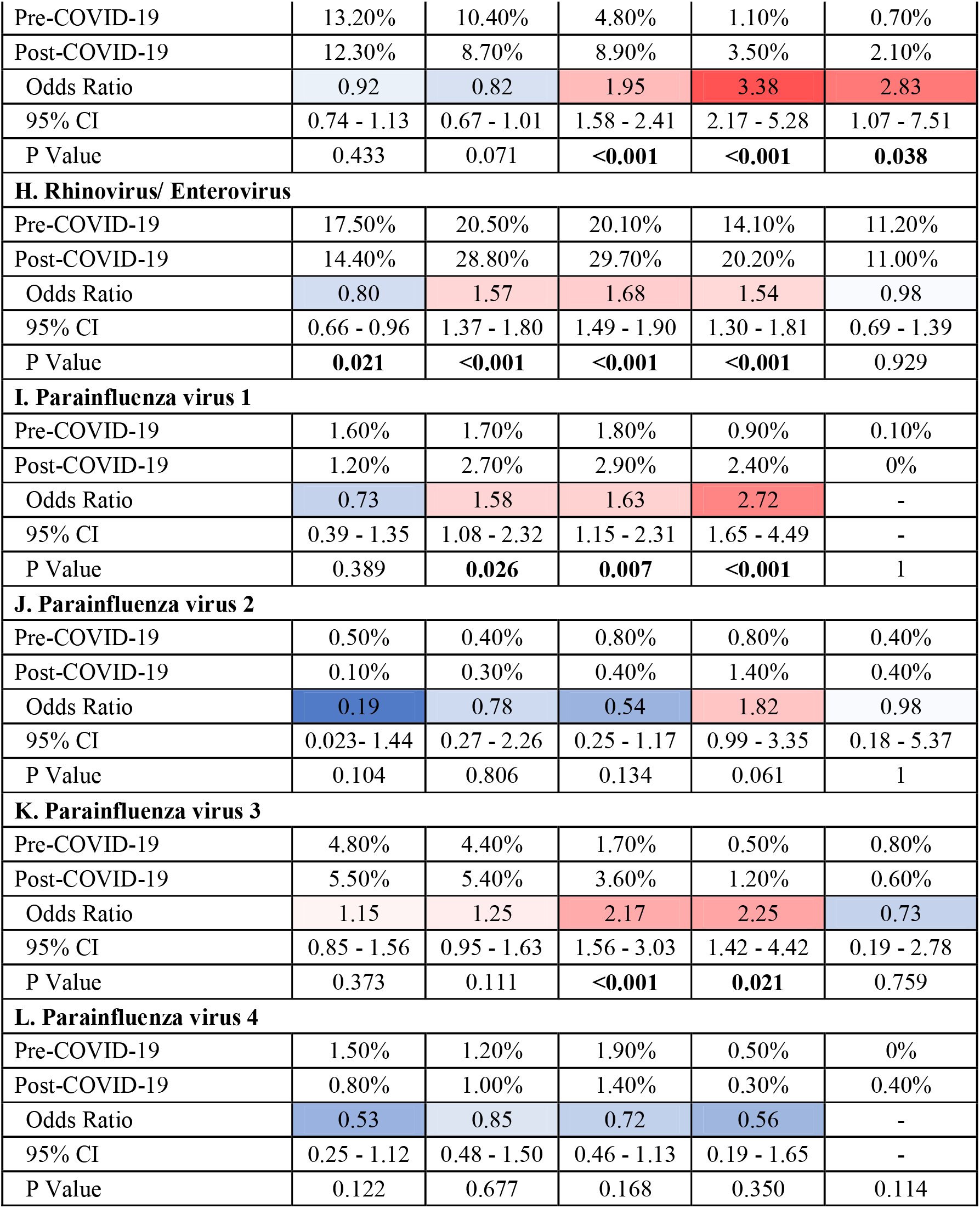
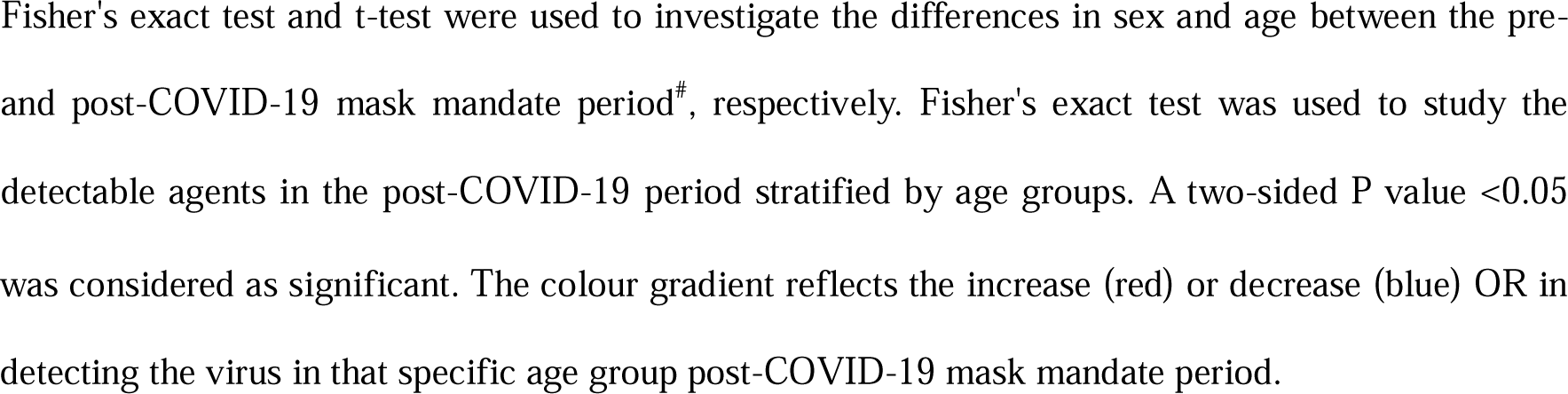
Demographics of study participants in the pre- and post-COVID-19 periods and the odd ratios of detectable agents in the post-COVID-19 period without and with age stratification.

**Figure 1.**
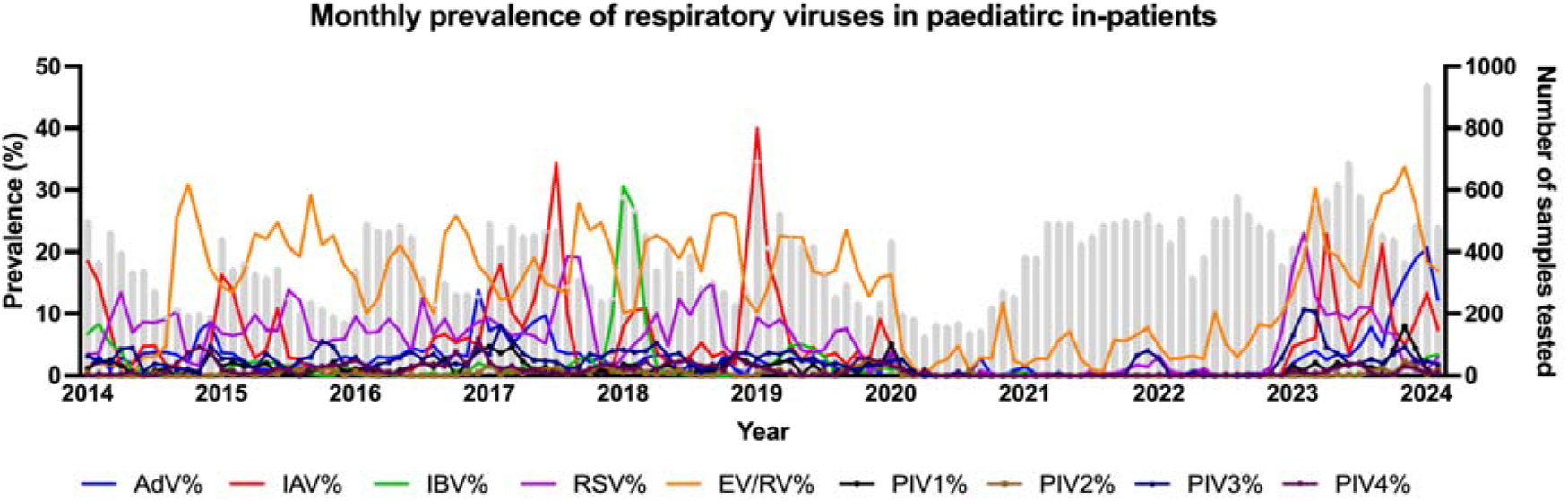
Prevalence of respiratory virus from paediatric inpatients before and after the SARS-CoV-2 pandemic. Monthly prevalence of detected respiratory viruses (left y-axis) with respective number of tested samples (columns in grey depicted by the right y-axis) from January 2015 to February 2024. AdV =adenovirus; IVs = influenza viruses; EV/RV = enterovirus/rhinovirus; RSV = respiratory syncytial viruses; PIVs = parainfluenza viruses.

The mean age of hospitalized patients increased from 3.49y ± 0.03y before COVID-19 to 4.37y ± 0.05y after COVID-19 mask mandate (P<0.001, Table 1). Generally, the rate of single virus infection was significantly higher in the post-COVID-19 mask mandate period (48.40% vs 43.00%, P<0.01) (**Table 1**). The age-groups stratified analysis revealed that AdV had a remarkably escalated odds ratio for all age groups (OR: 4.53, 4.03, 2.35, 2.46, 1.31); meanwhile, the odds ratio decreased as the age increased. In addition, RSV had a staggering increase in odds ratio in older children, especially in 6y to <12y (OR: 3.38, P<0.01), despite RSV typically contributes to hospitalization of children below five years old before the pandemic (2).

More importantly, the odds ratio of having more than one virus detected was higher, particularly in the age groups 1y to <3y (OR: 2.97, P<0.01), 3y to <6y (OR: 3.05, P<0.01), and 6y to <12y (OR: 2.70, P<0.01). The cases (n=298) and the detection rate (4.96 of co-infection in the one-year post-COVID-19 mask mandate exceeded the average of the four-year pre-COVID-19 era (n=97, 2.07%, P<0.001). Before COVID-19, EV/RV accounted for 68.4% of all viral co-infections, followed by RSV (35.4%), IAV (25.3%) and AdV (22.7%). Whereas in the post-COVID-19 mask mandate period, EV/RV was still the predominant cause of viral co-infections (75.2%), followed by the outthrusting of AdV (52.3%), followed by RSV (27.9%) and IAV (18.8%).

In conclusion, our findings suggest that the public health measures to contain COVID-19 may have unintended consequences on the natural exposure and immunity of children to other respiratory viruses, which could increase their morbidity and mortality in the post-pandemic era. Our study reported a rebound of respiratory viruses, and increased multiple virus infections, after lifting the COVID-19 restrictions. We showed the changing patterns in AdV and RSV infection, which respectively affected young and older children. Most of the increased vulnerability to respiratory viruses in children was seen in patients aged 3y to <6y. This could be attributed to the lack of exposure to viruses during lockdown, which is essential to building up immunity in early childhood (3, 4). The increase in virus detection post-COVID-19 mask mandate may also contribute by the immune suppression by the prior SARS-CoV-2 infection in children. However, the causation is undiscernible as COVID-19 infection in children are mostly asymptomatic and do not require hospitalization (5). Therefore, it is important to monitor the trends and patterns of respiratory virus circulation and co-infection among children, especially those with underlying conditions or immunocompromised status, and to provide timely and appropriate preventive and therapeutic interventions, as the increase in co-detection of viruses may indicate worse clinical outcomes (6). Further studies are needed to elucidate the causal relationship between SARS-CoV-2 exposure and subsequent respiratory virus susceptibility, as well as the long-term effects of interrupted virus exposure on children’s immune development.

## Data Availability

All data produced in the present work are contained in the manuscript

## Funding source

This research did not receive any specific grant from funding agencies in the public, commercial, or not-for-profit sectors.

## Conflict of Interest Disclosure

The authors declare no conflict of interest.

## Declaration of Competing Interest

The authors declare that they have no known competing financial interests or personal relationships that could have appeared to influence the work reported in this paper.

## Declaration of generative AI and AI-assisted technologies in the writing process

During the preparation of this work the authors used Grammarly in order to perform grammar and spell check. After using this tool/service, the authors reviewed and edited the content as needed and take full responsibility for the content of the publication.

## Notes

### Competing Interest Statement

The authors have declared no competing interest.

### Funding Statement

No external funding was received

### Author Declarations

Clincial Research Ethics Committee (CREC) of The Joint The Chinese University of Hong Kong (CUHK) and New Territories East Cluster (NTEC) gave ethical approval Ref: 2019.120 and 2022.612 for this work

